# Comparison of four intensive care scores in prediction of outcome after Veno-Arterial ECMO: A single center retrospective study

**DOI:** 10.1101/2024.08.12.24311770

**Authors:** Suraj Sudarsanan, Praveen Sivadasan, Prem Chandra, Amr S Omar, Kathy Lynn Gaviola Atuel, Hafeez Ulla Lone, Hany O Ragab, Irshad Ehsan, Cornelia S Carr, Abdulrasheed Pattath, Abdulaziz Al khulaifi, Yasser Mahfouz Eltokhy Shouman, A/Wahid Mahmoud A.A. Al Mulla

## Abstract

**Background:** Assess the ability of APACHE II (acute physiology and chronic health evaluation II), SOFA (sequential organ failure assessment scores), Cardiac Surgery Score (CASUS) and SAVE (Survival after VA-ECMO) to predict outcomes in a cohort of patients undergoing Veno-Arterial ECMO (VA-ECMO).

**Methods:** Observational retrospective study of all patients admitted to Cardiothoracic Intensive Care Unit (CTICU) for a minimum duration of 24 hours after undergoing VA-ECMO insertion between years 2015 to 2022. Scores for APACHE II, SOFA and CASUS were calculated at 24 after ICU admission. SAVE score was calculated from the last available patient details within 24 hours of ECMO insertion. Demographic, clinical, and laboratory data relevant for the study was retrieved from electronic patient records.

**Results:** Pre-ECMO serum levels of lactates and creatinine were significantly associated with mortality. Lower ECMO flow rates at 4 hours and 12 hours after ECMO cannulation was significantly associated with survival to discharge. Development of arrythmias, acute kidney injury (AKI) and need of continuous renal replacement therapy (CRRT) while on ECMO were significantly associated with mortality. The APACHE-II, SOFA and CASUS, calculated at 24 hours of ICU admission were significantly higher amongst non-survivors. Following categorization of risk scores using ROC curve analysis, it was found that APACHE-II, SOFA and CASUS calculated at 24 hours of ICU admission after ECMO insertion demonstrated moderate predictive ability for mortality whereas SAVE score failed to predict mortality. APACHE-II >27 (AUC of 0.66) calculated at 24 hours of ICU admission after ECMO insertion, demonstrated the greatest predictive ability, for mortality. Multivariate logistic regression analysis of the four scores showed that APACHE-II > 27 and SOFA > 14 calculated at 24 hours of ICU admission after ECMO insertion, were independently significantly predictive of mortality.

**Conclusions:** The APACHE-II, SOFA and CASUS, calculated at 24 hours of ICU admission were significantly higher amongst non-survivors as compared to survivors. APACHE-II demonstrated the best mortality predictive ability. APACHE-II scores of 27 or above, and SOFA of 14 or above at 24 hours of ICU admission after ECMO cannulation can predict mortality and will aid physicians in decision making.

## Introduction

Veno-arterial extracorporeal membrane oxygenation (VA-ECMO) as part of Extracorporeal life support (ECLS) is a standard therapy for short-term hemodynamic support, in the setting of cardiogenic shock (CS), especially in progressively deteriorating patients, despite maximal medical therapy. VA-ECMO serves as a short-term ventricular assist device for Mechanical Circulatory support (MCS) that can be expeditiously placed at the bedside, in the emergency room, intensive care unit, cardiac catheterization suite, or operating room. This can be used as a bridge to recovery or as a bridge to decision providing clinicians the opportunity for further treatment and evaluation. (1,2). VA-ECMO has, of late, become popular for extracorporeal cardio-pulmonary resuscitation (ECPR), where it maintains vital organ perfusion during and immediately after cardiac arrest (3) Post-cardiotomy VA-ECMO is used when weaning from cardiopulmonary bypass (CPB) is difficult and it allows time for the ventricle to recover after complex surgeries. It is often electively instituted after heart transplantation as a bridge to recovery.

Prognosis and outcomes after the institution of VA-ECMO are largely dependent on the underlying indication, patient comorbidities, severity of organ dysfunction at initiation, and complications or adverse events during MCS. (4,5) Although it has life-saving potential, VA-ECMO is associated with complications especially vascular complications from cannulation. Apart from this, a patient on VA-ECMO is liable to develop neurologic injury, renal failure, liver failure, and sepsis as a sequalae of post-cardiogenic shock or cardiac arrest. (6,7). Despite notable advances in the quality of devices and in intensive care management of these patients, ECLS therapy is still associated with a high rate of mortality and complications (1).

Inappropriate and un-indicated application of VA-ECMO can lead to unnecessary prolongation of patient and family suffering with significant costs and resource utilization. (6,7). Due to these reasons, early prognostication assumes importance in patients undergoing treatment with VA-ECMO. Many survival prediction models are available for critically ill patients admitted in Intensive Care Units (ICU). These scores however have not demonstrated consistent results when applied to patients on ECLS (8–11).

Only a few such models, specifically designed for VA-ECMO are available. Schmidt et al. created the SAVE scoring system utilizing the Extracorporeal Life Support Organization (ELSO) registry to predict survival to discharge in VA-ECMO patients (12). Chen et al combined serum lactate to SAVE score and created Modified SAVE score for outcome prediction for patients who underwent urgent VA-ECMO in the emergency department (13). The ENCOURAGE score was created utilizing a bi-institutional database and predicts survival to ICU discharge in VA-ECMO patients (14). The REMEMBER score comprising 6 pre-ECMO variables, was designed to predict in-hospital mortality for patients on VA-ECMO after isolated CABG for refractory cardiogenic shock. (15).

We describe our experience with comparison of the predictive accuracy of four prediction models in predicting the outcome in a cohort of patients undergoing institution of VA-ECMO for cardiogenic shock or cardiac arrest. We tested two general ICU risk scores: APACHE II (acute physiology and chronic health evaluation II) and SOFA (sequential organ failure assessment), one score specific to cardiac surgery patients: CASUS (Cardiac surgery risk score) and one score specific to VA-ECMO: SAVE (survival after veno-arterial-ECMO).

## Materials and Methods

This was an observational retrospective study, conducted in the cardiothoracic intensive care unit (CTICU) of a tertiary cardiac center, of all patients admitted to CTICU after undergoing VA-ECMO insertion, between years 2015 to 2022. Patients whose ICU course, post-ECMO insertion, was less than 24 hours and patients who after ECMO insertion were not admitted to the CTICU were excluded from the study. The research study was approved by the Institutional Review Board of our institution (study protocol # MRC-01-22-701), which waived informed consent as this was a retrospective review of existing patient records without revealing any identity of participants.

The authors recorded the demographic, clinical, and laboratory data relevant for the study from the electronic patient records (Cerner Millennium need country, and Dendrite Clinical Systems, London, United Kingdom). This included pre-ECMO patient demographics, clinical, laboratory, hemodynamic parameters (pre-ECMO as well as while on ECMO), cannulation details (location, type, constitution), procedures done while on ECMO (revascularization-surgical or percutaneous, re-look angiography, definitive cardiac surgical procedures), complications on ECMO and outcome. Pre-ECMO variables were the last values available within 24 hours of cannulation.

Indications and contra-indications for insertion of VA-ECMO were based on local protocol and ELSO guidelines. The VA-ECMO pumps used were either CARDIOHELP (Maquet, Wayne, NJ) with HLS Set Advanced Oxygenator (Maquet Cardiopulmonary GmbH, Rastatt) or CentriMag (Levitronix, Waltham, MA) with Medos Hilite Oxygenator (MEDOS Medizintechnik AG, Stolberg). Cannulation strategies depended upon clinical scenario and surgeon’s preference. Arterial cannulation was established via femoral, axillary, subclavian or direct aortic route, whereas venous cannulation was usually established via femoral, internal jugular or central right atrial routes. Some patients with concomitant poor lung function needed reconfiguration from VA-to Veno-arterio-venous extracorporeal membrane oxygenation (VAV-ECMO) to combat hypoxia.

Patients were cannulated bedside; percutaneous femoral cannulation was commonly used in cases of emergency cannulations in the emergency room, cardiac catheterization suites or in-patient wards following cardiac arrest as ECPR. Central cannulation via aortic and right atrial route was performed in the operating room in selected cases of post-cardiotomy shock requiring VA-ECMO support. Subclavian or axillary route for arterial cannulation was utilized when femoral artery was deemed unusable due to injury, atherosclerosis, small sized lumen etc. With all femoral arterial cannulations, an ipsilateral antegrade limb perfusion cannula was placed (percutaneously or by surgical cutdown) to prevent ischemic complications, as mandated by institutional protocol. Patients were anticoagulated with Heparin infusion to maintain an activated partial thromboplastin time (aPTT) between 40-60 seconds during the entire course of ECMO. Heparin infusion was kept on hold when there was excessive bleeding and /or coagulopathy and in such cases the duration of cessation of heparin was reviewed during the twice daily multi-disciplinary clinical rounds.

Scores for APACHE II, SOFA and CASUS were calculated at 24 after ICU admission. SAVE score was calculated from the last available patient details within 24 hours of ECMO insertion. For calculation of SAVE score in ECPR patients, presenting to the emergency room with cardiac arrest, we used the last available patient details in electronic record or assumed to have normal parameters if no previous records could be found.

### Statistical analysis

Descriptive statistics were used to summarize and determine the sample characteristics and distribution of participants’ data. The normally distributed data and results were reported with mean and standard deviation (SD), whereas median and inter-quartile range (IQR) were used for skewed data distribution. Categorical data were summarized using frequencies and proportions.

APACHE-II, SOFA, SAVE, and CASUS scores were calculated for all patients. The main focus of the data analysis in our current research study was to determine the predictive accuracy of these four scores APACHE-II, SOFA, SAVE, and CASUS in predicting non-survivors in a cohort of patients undergoing insertion of VA-ECMO for cardiogenic shock or cardiac arrest.

Associations between two or more qualitative variables were assessed using Chi-square (χ2) test or Fisher Exact test as appropriate. Quantitative data between the two independent groups (survivors and non-survivors) were analyzed using unpaired t or Mann Whitney U test as appropriate. Univariate and multivariate logistic regression methods were applied to assess the predictive values of each score along with other potential confounders and predictors associated with non-survivors (yes/no). For multivariate regression models, variables were considered, if statistically significant at the P <0 .10 level in univariate analysis or if determined to be clinically important. Discriminative ability of the logistic regression model was assessed by area under the receiver-operating characteristics curve (ROC).

Sensitivity, specificity, positive and negative predictive values, and likelihood ratios of these parameters were calculated. ROC curve was constructed, and indices calculated to derive best suitable cut-off values for the above four risk scores to assess model discrimination and predictive accuracy. Pictorial presentations of the key results were made using appropriate statistical graphs. All P values presented were two-tailed, and P values <0.05 was considered as statistically significant. All Statistical analyses will be done using statistical packages SPSS version 29.0 (Armonk, NY: IBM Corp) and Epi Info 2000 (Centers for Disease Control and Prevention, Atlanta, GA).

## Results

Between the years 2015 and 2022, all patients who underwent VA-ECMO insertion in our institution for cardiogenic shock or cardiac arrest were screened and 142 patients who were managed in CTICU after ECMO insertion for a minimum of 24 hours were included in the study. Mean age of the study group was 49.89 ± 11.70 years. Males vastly outnumbered females in the study group (88.73%) and the study group had a mixed ethnic/racial profile The indications for VA-ECMO insertion in the study group included ECPR in 50%, post cardiotomy shock following cardiac surgery in 29%, cardiogenic shock in 17.5% and other etiologies in 3.5% cases. Peripheral cannulation (femero-femoral and Femero-axillary/subclavian) was employed in 95% cases and central cannulation in 5% cases. Intra-aortic balloon pump (IABP) was concomitantly present in 76% of patients. 10% of patients need reconfiguration of VA-ECMO to-Veno-arterio-venous extra corporal membrane oxygenation (VAV ECMO Outcome assessment of the study group (Table 2) showed that of the 142 patients included, 120 patients could be weaned from VA-ECMO (84%), but only 59 patients were discharged alive from hospital after ECMO insertion (41.51%), whereas 83 patients died while in hospital (58.52%). Of the patients who died in hospital, 22 (26.52%) died while on ECMO, 47 (56.62%) died in the ICU after ECMO removal and 14 (16.86%) died after transfer from ICU to the floor (Table 4). The absolute percentage of mortality across various cut-offs scores for each risk score presented in Figure 4, indicated higher the values of risk scores cut-off associated with higher percentage of mortality On assessment of quantitative predictors and their association with survivors and non-survivors is shown in Table 1.a. Amongst the pre-ECMO variables recorded, only lactates and creatinine levels showed statistically significant elevation in non-survivors as compared to survivors (P<0.05). This study observed that ECMO flow at 4 hours post ECMO insertion was significantly higher amongst non-survivors as compared to survivors (3.24 ± 0.78 L/min vs 3.0 ± 0.60 L/min, P=0.05), the same trend was found with ECMO flows at 12 hours post ECMO insertion (3.17± 0.89 L/min vs 2.89 ± 0.67 L/min, P=0.03). The attainment of lower flows in survivors at 4 hours and 12 hours post-ECMO signifies recovery of LV function. The length of hospital stay (LOH) was significantly higher in survivors as compared to non-survivors (median 40; inter-quartile range (IQR) 18,79 days vs median 14; IQR 6,34 days; P=0.007).

**Table 1.a:** Quantitative predictors and their association with survivors and non-survivors.

| Parameters | Non-survivors<br>(n=83)<br>mean ± SD /<br>Median (IQR) | Survivors<br>(n=59)<br>mean ± SD /<br>Median (IQR) | Unadjusted<br>OR (95% CI) | P-value |
| --- | --- | --- | --- | --- |
| Age (years) | 50.47 ± 12 | 49.07 ± 11.30 | 1.01(0.98,1.04) | 0.48 |
| BMI (kg/m<sup>2</sup>) | 26.82 ± 5.32 | 26.05 ± 4.61 | 1.03(0.96,1.10) | 0.36 |
| EF (%) | 35.42 ± 17 | 35.55 ± 16.42 | 1.01(0.97,1) | 0.96 |
| ECMO Duration<br>(hours) | 125(125,193) | 116 (51,170) | 1.01(0.99,1) | 0.14 |
| **Pre-ECMO** |  |  |  |  |
| Low Flow Time<br>(minutes) | 19 (2,35) | 10 (0,30) | 1.01(0.99,1) | 0.57 |
| Ventilation (hours) | 3 (1,10) | 2 (1,6) | 1.01(0.99,1) | 0.93 |
| Lactate (mmol/L) | 7.2 (3.32,13.91) | 5.9 (3.91,10) | 1.02(0.99,1.12) | **0.07** |
| WBC (X 10<sup>9</sup>/L) | 14.25 ± 6.28 | 13.48 ± 6.87 | 1.01(0.96,1) | 0.50 |
| Platelets (X 10<sup>9</sup>/L) | 224 (131.5,312.5) | 203.5(135.6,265.3) | 1.01(0.99,1) | 0.60 |
| Bilirubin<br>(micromol/L) | 13 (7, 36.75) | 11.95(6.55,28.75) | 1.02(0.99,1) | 0.27 |
| Creatinine<br>(micromol/L) | 143(100.51,240.52) | 102 (79.25,134.53) | 1.01(0.99,1) | **0.07** |
| ALT (Units/L) | 69 (24,240.52) | 33 (20,136) | 1.01(0.99,1) | 0.55 |
| **On ECMO** |  |  |  |  |
| ECMO flow(L/min)<br>@4 hrs | 3.24 ± 0.78 | 3.0 ± 0.60 | 1.6 (0.99,2.74) | **0.05** |
| ECMO flow (L/min)<br>@ 12 hrs | 3.17 ± 0.89 | 2.89 ± 0.67 | 1.6 (1, 2.72) | **0.03** |
| LOV (hours) | 216 (96,360) | 255 (150,528) | 1.02(0.99,1) | 0.41 |
| LOS-ICU (days) | 12 (5,23) | 16 (11,30) | 1.01(0.99,1) | 0.67 |
| LOS-Hospital (days) | 14 (6,34) | 40 (18,79) | 0.98(0.97,0.99) | **0.007** |
| **Risk Scores** |  |  |  |  |
| CASUS | 20.45 ± 4.15 | 18.24 ± 4.11 | 1.15(1.04,1.26) | **0.003** |
| APACHE-II | 30.13 ± 6.53 | 26.24 ± 6.82 | 1.09(1.03,1.16) | **0.001** |
| SOFA | 16.22 ± 3.50 | 14.05 ± 3.81 | 1.20(1.07,1.34) | **0.001** |
| SAVE | -5.86 ± 5.62 | -4.49 ± 6.12 | 0.96(0.90,1.01) | 0.16 |

CI: Confidence interval; OR: Odds ratio.

Univariate logistic regression analysis of categorical predictors and their association with non-survivors has been depicted in Table 1b. Patients who developed acute kidney injury (AKI) on ECMO were significantly associated with higher risk of mortality as compared to those who didn’t develop AKI (unadjusted odds ratio (OR) 4.22, 95% CI 1.75, 10.14, P=0.001). Our study relied on Acute Kidney Injury Network (AKIN) criteria for diagnosing AKI. Similarly, patients who underwent continuous renal replacement therapy (CRRT) had a significantly higher mortality than those who did not (unadjusted OR 2.99, 95% CI 1.41, 6.34, P=0.004). The study also revealed that patients who developed arrythmias while on ECMO [ventricular tachycardia (VT), ventricular fibrillation (VF) or atrial fibrillation (AF)] had a significantly higher mortality as compared to those who did not (66.21% vs 50%, P=0.051). Mortality of patients who were put on ECMO following ECPR was higher compared to those put on ECMO following cardiogenic shock and post-cardiotomy (62% vs 52% and 51.2% respectively), but this was not statistically significant.

**Table 1.b:** Categorical Predictors and their association with non-survivors (Univariate Logistic regression analysis).

| Predictors | Non-survivors<br>n/N (%) | Unadjusted<br>OR (95% CI) | P-value |
| --- | --- | --- | --- |
| **Gender** |  |  |  |
| Male | 74/126 (58.71%) | 1 (reference) | 0.850 |
| Female | 9/16 (56.32%) | 0.90 (0.31,2.58) |  |
| **Ethnicity** |  |  |  |
| Indian | 54/92 (58.72%) | 1 (reference) |  |
| Arab | 20/35 (57.11%) | 0.93(0.42,2.06) | 0.874 |
| Asian | 3/4 (75%) | 2.11(0.21,21.06) | 0.524 |
| Others | 6/11 (54.52%) | 0.84(0.24,2.96) | 0.792 |
| **Cardiac arrest location** |  |  |  |
| No Cardiac arrest | 26/50 (52%) | 1 (reference) |  |
| In hospital | 52/87 (59.83%) | 1.37(0.68,2.76) | 0.370 |
| Out of Hospital | 5/5 (100%) | - | 0.990 |
| **Indication for ECMO** |  |  |  |
| ECPR | 44/71 (62%) | 1 (reference) |  |
| Cardiogenic shock | 13/25 (52%) | 0.66(0.26,1.66) | 0.381 |
| Post-cardiotomy shock | 21/41(51.22%) | 0.64(0.29,1.40) | 0.262 |
| Others | 5/5 (100%) | - | 0.990 |
| **Location of cannulation** |  |  |  |
| Operating Room | 22/44 (50%) | 1 (reference) |  |
| Cath-Lab | 32/49 (65.3%) | 1.88(0.81,4.33) | 0.137 |
| ICU | 23/35 (65.70%) | 1.91(0.76,4.78) | 0.163 |
| ER | 5/11 (45.51%) | 0.83(0.22,3.13) | 0.788 |
| HDU/Floor | 1 /3(33.33%) | 0.5 (0.04,5.92) | 0.583 |
| **Surgery while on ECMO** |  |  |  |
| No Surgery | 49/80 (61.32%) | 1 (reference) |  |
| CABG | 19/37 (51.41%) | 0.66(0.30,1.46) | 0.312 |
| Valve Surgery | 7/9 (77.82 %) | 2.21(0.43,11.35) | 0.340 |
| CABG + Valve | 2/5 (40%) | 0.42(0.06,2.66) | 0.351 |
| Others | 6/11 (54.51%) | 1.05(0.23,4.72) | 0.940 |

**Table 1.b: Categorical Predictors and their association with non-survivors (Univariate Logistic regression analysis) ...contd**
| Predictors | Non-survivors<br>n/N (%) | Unadjusted<br>OR (95% CI) | P-value |
| --- | --- | --- | --- |
| **Type of MI preceding ECMO** |  |  |  |
| No MI | 18/33 (54.51%) | 1 (reference) |  |
| NSTEMI | 24/41 (58.52%) | 1.17(0.47,2.96) | 0.731 |
| STEMI | 41/68 (60.30%) | 1.26(0.54,2.93) | 0.580 |
| **Type of VA-ECMO** |  |  |  |
| Femero-femoral | 72 (86.72%) | 1 (reference) |  |
| Femro- Axillary/Subclavian | 5/6 (83.34%) | 0.26(0.10,3.65) | 0.321 |
| Central | 6/7 (85.72%) | 0.26(0.03,2.30) | 0.220 |
| **Miscellaneous variables** |  |  |  |
| **IABP on ECMO** |  |  |  |
| No | 18/34 (52.94%) | 1 (reference) |  |
| Yes | 65/108 (60.21%) | 1.34(0.61,2.91) | 0.456 |
| **Conversion to V-AV** |  |  |  |
| No | 72/128 (56.25%) | 1 (reference) |  |
| Yes | 11/14 (78.63%) | 2.85(0.76,10.71) | 0.121 |
| **Additional ECMO runs** |  |  |  |
| No | 73/128 (57%) | 1 (reference) |  |
| Yes | 10/14 (71.42%) | 1.88(0.56,6.32) | 0.306 |
| **Arrhythmias on ECMO** |  |  |  |
| No | 34/68 (50%) | 1 (reference) |  |
| Yes | 49/74 (66.21%) | 1.96(0.99,3.86) | 0.051 |
| **Acute Kidney Injury** |  |  |  |
| No | 9/29(31.03%) | 1 (reference) |  |
| Yes | 74/113 (65.50%) | 4.22(1.75,10.14) | **0.001** |
| **CRRT on ECMO** |  |  |  |
| No | 45/91 (49.45%) | 1 (reference) |  |
| Yes | 38 /51(74.53%) | 2.99(1.41,6.34) | **0.004** |

**Table 1.b: Categorical Predictors and their association with non-survivors (Univariate Logistic regression analysis) ...contd**
| Predictors | Non-survivors<br>n/N (%) | Unadjusted<br>OR (95% CI) | P-value |
| --- | --- | --- | --- |
| Open chest on ECMO |  |  |  |
| No | 27/40 (67.5%) | 1 (reference) |  |
| Yes | 56/102 (55%) | 0.59 (0.27, 1.26) | 0.173 |
| ICU readmission post ECMO |  |  |  |
| No | 69/115 (60%) | 1 (reference) |  |
| Yes | 14/27 (51.9%) | 0.72(0.31,1.67) | 0.441 |
| Complications on ECMO |  |  |  |
| No | 4/9 (44.4%) | 1 (reference) |  |
| Yes | 79/133 (59.4%) | 1.83(0.47,7.12) | 0.384 |
| **Risk Score** |  |  |  |
| CASUS ≤ 18 | 33/63 (52.41%) | 1 (reference) | 0.190 |
| CASUS > 18 | 50/79 (63.32%) | 1.56(0.79,3.07) |  |
| APACHE-II ≤27 | 26/61 (42.62%) | 1 (reference) | **0.001** |
| APACHE-II >27 | 57/81 (70.40%) | 3.19(1.59,6.41) |  |
| SOFA ≤ 14 | 22/56 (39.3%) | 1 (reference) | **<0.001** |
| SOFA >14 | 61/86 (71%) | 3.77(1.85,7.67) |  |
| SAVE ≤ -6 | 44/73 (60.31%) | 1 (reference) | 0.650 |
| SAVE > -6 | 39/69 (56.55%) | 0.85(0.43,1.67) |  |

*CI: Confidence interval; OR: Odds ratio* *'n' is the total number of non-survivors cases whereas 'N' is the total number of participants included against* *each specific variable/parameter*

**Table 2:** Outcome characteristics of study population.

| Disposition | Frequency (%) n= 142 |
| --- | --- |
| Successful weaning from VA-ECMO | 120 (84.51%) |
| Hospital Discharge alive | 59 (41.51%) |
| Died in Hospital | 83 (58.52%) |
| Death Disposition | Frequency (5) n=83 |
| Died on ECMO | 22 (26.52%) |
| Died after ECMO removal in ICU | 47 (56.62%) |
| Died after ICU discharge | 14 (16.86%) |

As shown in Table 1.a, amongst the 4 risk scores compared in the study, SAVE score was not significantly different amongst survivors and non-survivors. The value of the other three scores, calculated at 24 hours of ICU admission were significantly higher amongst non-survivors as compared to survivors (CASUS 20.45 ± 4.15 vs 18.24 ± 4.11; P=0.003), APACHE-II 30.13±6.53 vs 26.24 ± 6.82, P=0.001), SOFA (16.22 ± 3.50 vs 14.05 ± 3.81, P=0.001). Following categorization of risk scores using ROC curve analysis, it was found that among the four risk scores, APACHE-II >27 (unadjusted OR 3.19, 95% CI 1.59, 6.41, P=0.001) and SOFA>14 (unadjusted OR 3.77, 95% CI 1.85, 7.67, P<0.001) had significant association with higher risk of mortality.

We performed and explored two multivariate logistic regression analysis models, one including four independent risk scores, and other model included potential predictors and confounding factors in addition to four independent risk scores and the respective findings had been depicted in tables 3.a and 3.b. The results showed that, after adjustment for potential predictors and confounding factors, APACHE-II >27 (adjusted OR 2.61, 95% CI 1.06, 6.44, P=0.037) and SOFA>14 (adjusted OR 4.68, 95% CI 1.90, 11.55, P=0.001) were independently significantly predictive of mortality as shown in Table 3.b. Furthermore, we did evaluate the predictive accuracy of both multivariate regression models developed using ROC curve and indices; the results indicated both regression models have had almost similar discriminative ability of the models and were found to have good accuracies s (AUC 0.72, 95% CI 0.64, 0.80 vs 0.79, 95% CI 0.71, 0.86) shown in Figure 1. The distribution of these four risk scores across survivors and non-survivors depicted using Box plots, Figure 2.

**Table 3. a:** Predictive assessment of potential risk scores with non-survivors (Multivariate logistic regression analysis).

| **Risk Scores** | **Adjusted**<br>**Odds ratio**<br>**(aOR)** | **95% CI for aOR** | **P-value** |
| --- | --- | --- | --- |
| CASUS > 18 | 1.07 | 0.50, 2.32 | 0.857 |
| APACHE-II >27 | 2.74 | 1.23, 6.12 | **0.014** |
| SOFA >14 | 3.05 | 1.42, 6.52 | **0.004** |
| SAVE > -6 | 1.51 | 0.68, 3.36 | 0.311 |

**Table 3. b:** Predictive assessment of risk scores with other specific potentially clinically relevant predictors with non-survivors (Multivariate logistic regression analysis).

| **Risk Scores/potential predictors** | **Adjusted Odds ratio (aOR)** | **95% CI for aOR** | **P-value** |
| --- | --- | --- | --- |
| CASUS > 18 | 1.14 | 0.48, 2.74 | 0.765 |
| APACHE-II >27 | 2.61 | 1.06, 6.44 | 0.037 |
| SOFA >14 | 4.68 | 1.90, 11.55 | 0.001 |
| SAVE > -6 | 2.08 | 0.85, 5.08 | 0.107 |
| ECMO duration | 1.01 | 1.00, 1.01 | 0.013 |
| Low flow minutes | 1.00 | 1.00, 1.00 | 0.994 |
| **Indication** |  |  |  |
| ECPR | reference |  |  |
| Cardiogenic shock | 0.40 | 0.12, 1.30 | 0.126 |
| Others | 0.73 | 0.15, 3.67 | 0.705 |
| **Location of cannulation** |  |  |  |
| Operating Room | reference |  |  |
| Cath-Lab | 2.31 | 0.41, 12.88 | 0.341 |
| ICU | 1.83 | 0.40, 8.35 | 0.438 |
| ER | 1.21 | 0.14, 10.41 | 0.865 |
| HDU/Floor | 1.03 | 0.04, 27.55 | 0.985 |
| **Type of MI associated** |  |  |  |
| No MI | reference |  |  |
| NSTEMI | 1.82 | 0.61, 5.46 | 0.282 |
| STEMI | 1.64 | 0.57, 4.71 | 0.360 |
| Additional ECMO runs (yes) | 1.04 | 0.24, 4.58 | 0.957 |
| Definitive procedure on ECMO (yes) | 0.33 | 0.12, 0.90 | 0.031 |

*CASUS ≤ 18, APACHE-II ≤27, SOFA ≤ 14, SAVE ≤ -6 were considered as reference categories*

**Table 4:**
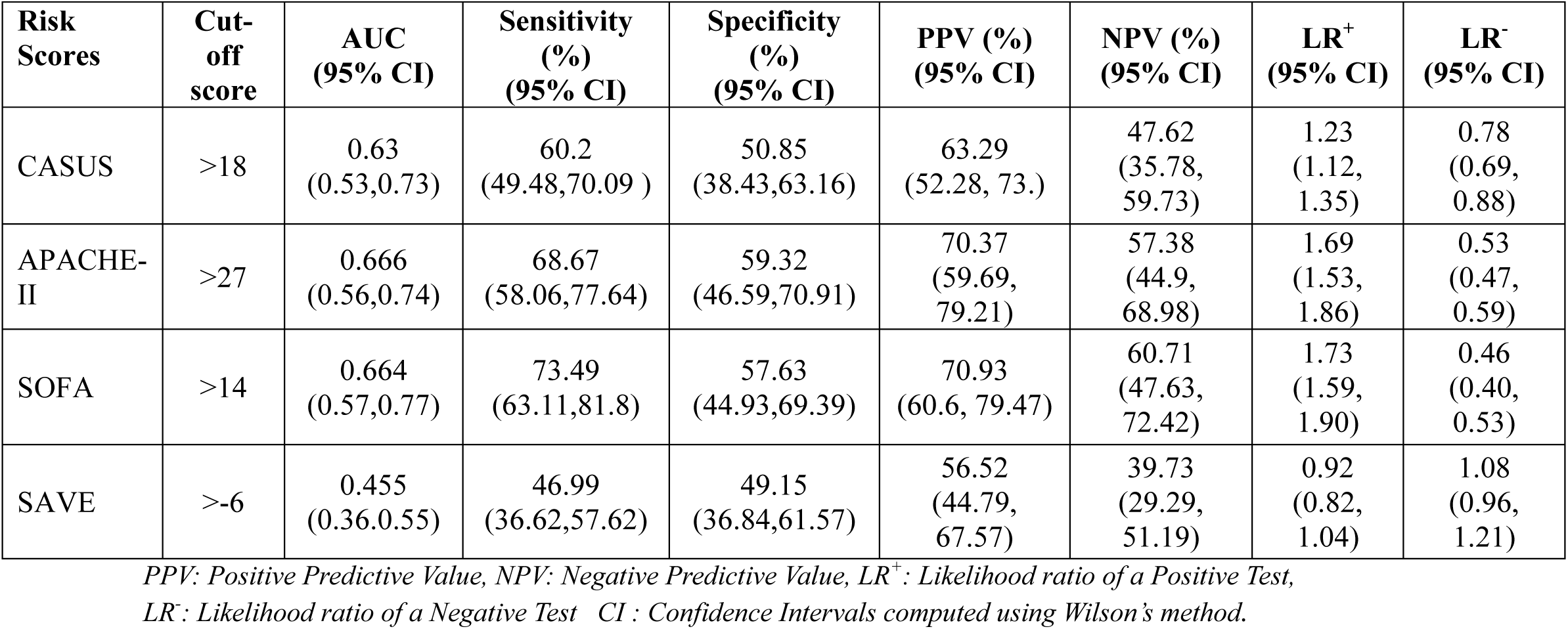
Diagnostic test evaluation of various cut-off values of Scores in predicting mortality.

| **Risk Scores** | **Cut-off score** | **AUC (95% CI)** | **Sensitivity (%) (95% CI)** | **Specificity (%) (95% CI)** | **PPV (%) (95% CI)** | **NPV (%) (95% CI)** | **LR<sup>+</sup> (95% CI)** | **LR<sup>-</sup> (95% CI)** |
| --- | --- | --- | --- | --- | --- | --- | --- | --- |
| CASUS | >18 | 0.63<br>(0.53,0.73) | 60.2<br>(49.48,70.09 ) | 50.85<br>(38.43,63.16) | 63.29<br>(52.28, 73.) | 47.62<br>(35.78, 59.73) | 1.23<br>(1.12, 1.35) | 0.78<br>(0.69, 0.88) |
| APACHE-II | >27 | 0.666<br>(0.56,0.74) | 68.67<br>(58.06,77.64) | 59.32<br>(46.59,70.91) | 70.37<br>(59.69, 79.21) | 57.38<br>(44.9, 68.98) | 1.69<br>(1.53, 1.86) | 0.53<br>(0.47, 0.59) |
| SOFA | >14 | 0.664<br>(0.57,0.77) | 73.49<br>(63.11,81.8) | 57.63<br>(44.93,69.39) | 70.93<br>(60.6, 79.47) | 60.71<br>(47.63, 72.42) | 1.73<br>(1.59, 1.90) | 0.46<br>(0.40, 0.53) |
| SAVE | >-6 | 0.455<br>(0.36,0.55) | 46.99<br>(36.62,57.62) | 49.15<br>(36.84,61.57) | 56.52<br>(44.79, 67.57) | 39.73<br>(29.29, 51.19) | 0.92<br>(0.82, 1.04) | 1.08<br>(0.96, 1.21) |

PPV: Positive Predictive Value, NPV: Negative Predictive Value, LR<sup>+</sup>: Likelihood ratio of a Positive Test,
LR<sup>-</sup>: Likelihood ratio of a Negative Test CI: Confidence Intervals computed using Wilson's method.

**Figure 1.**
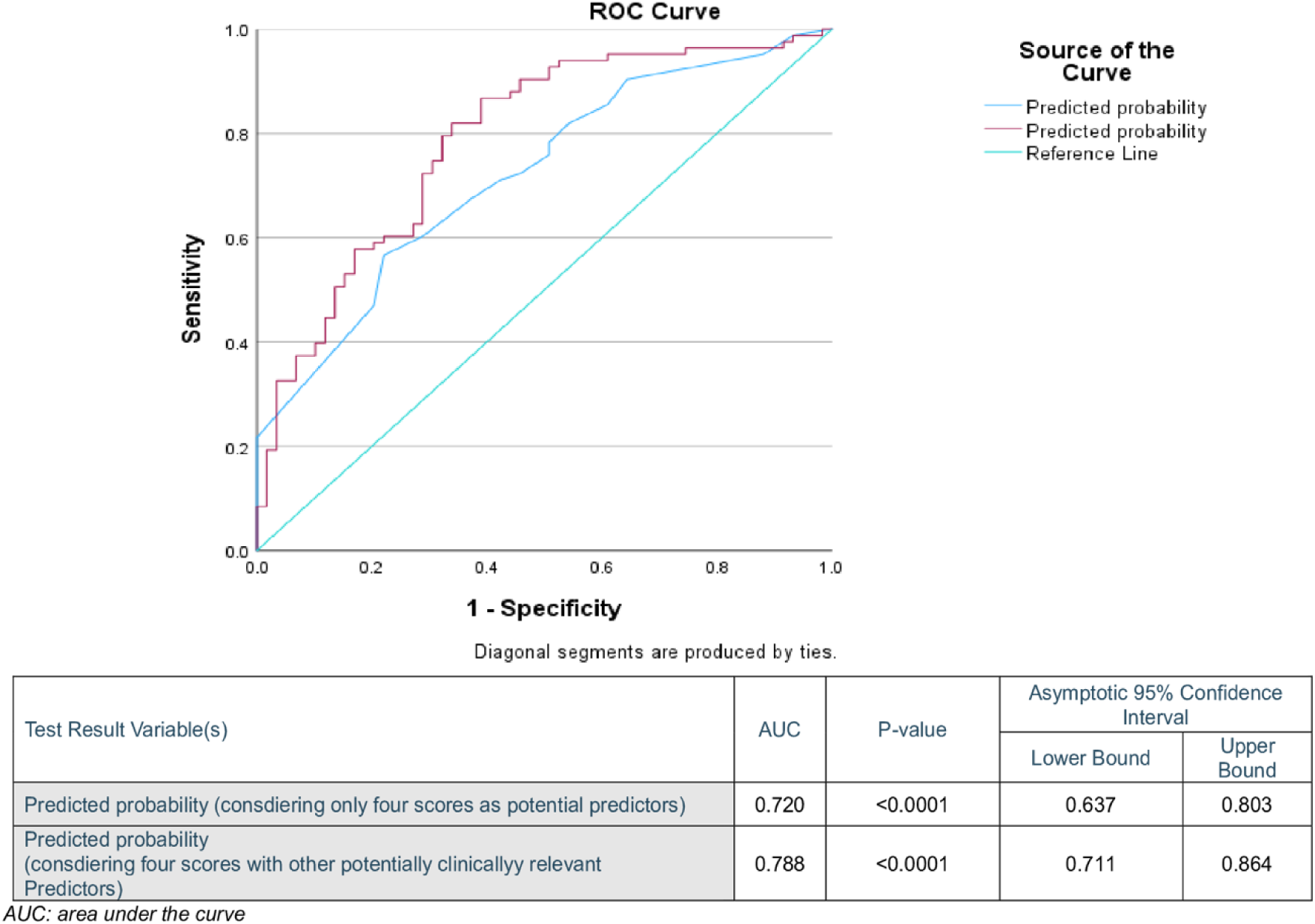
Predictive accuracy evaluation of both multivariate regression models using ROC curve and indicies.

**Figure 2.**
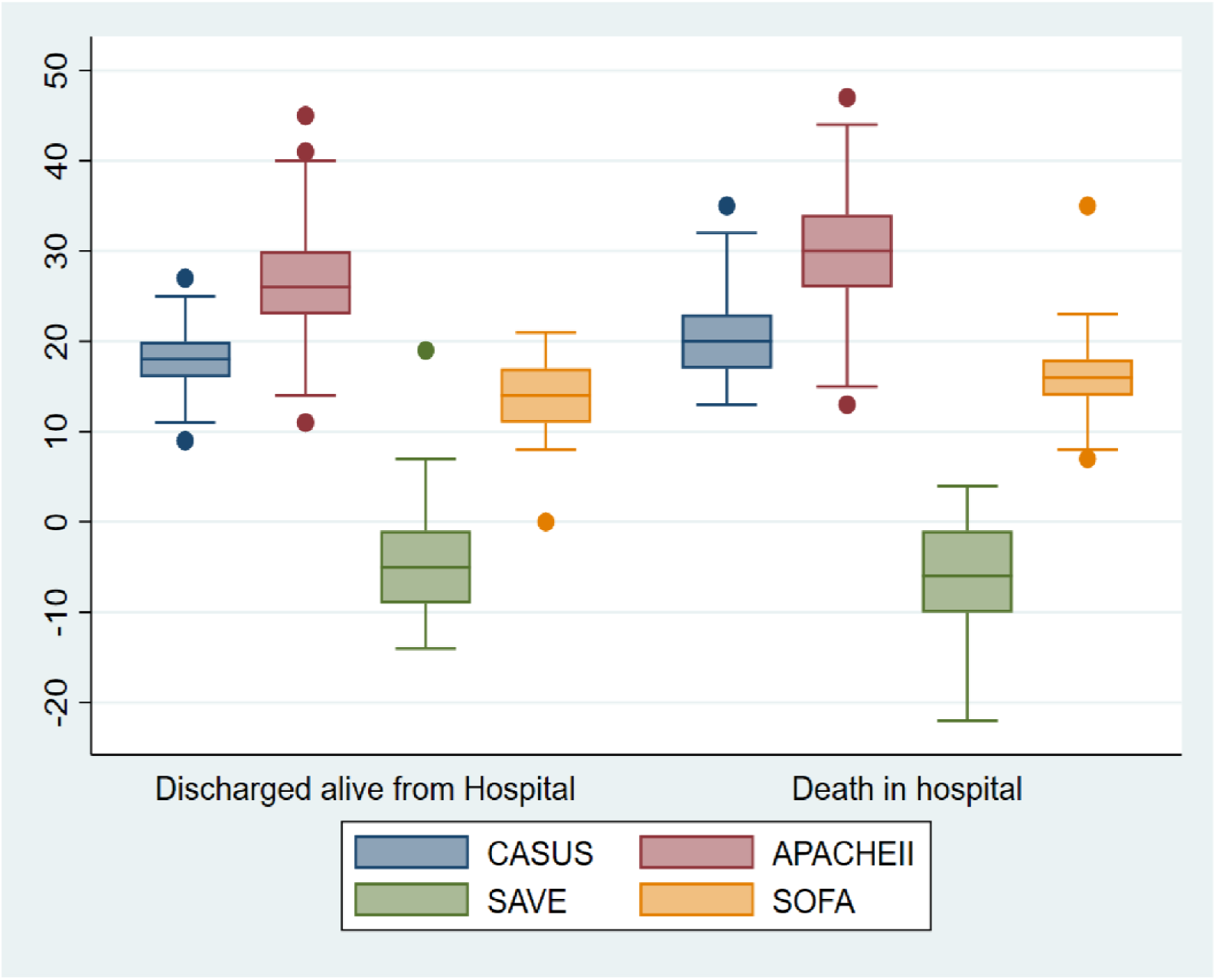
Box Plots depicting distribution of four risk scores in survivors and non-survivors.

ROC curves were plotted to determine an optimum cutoff value of these four risk scores in predicting mortality (Figure 3), the following cut-offs were identified to be associated with mortality, CASUS>18, APACHE-II>27, SOFA>14 and SAVE> −6. The results of diagnostic test evaluation using indices sensitivity, specificity, positive predictive value (PPV), negative predictive value (NPV), likelihood ratio of a positive test (LR+) and likelihood ratio of a negative test (LR-) of various cut-off values of these fours risk scores in predicting mortality are shown in Table 4.

**Figure 3.**
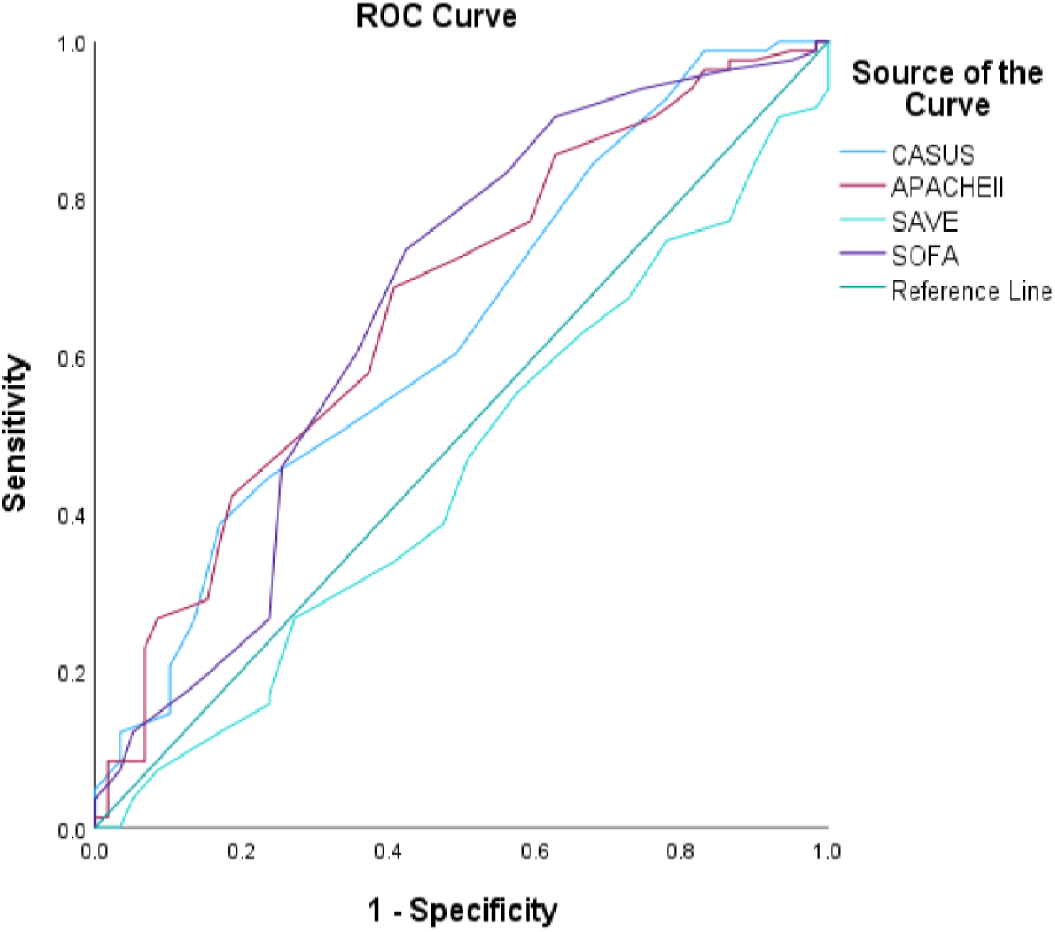
ROC curve to determine an optimum cutoff value for Scores in predicting mortality.

**Figure 4.**
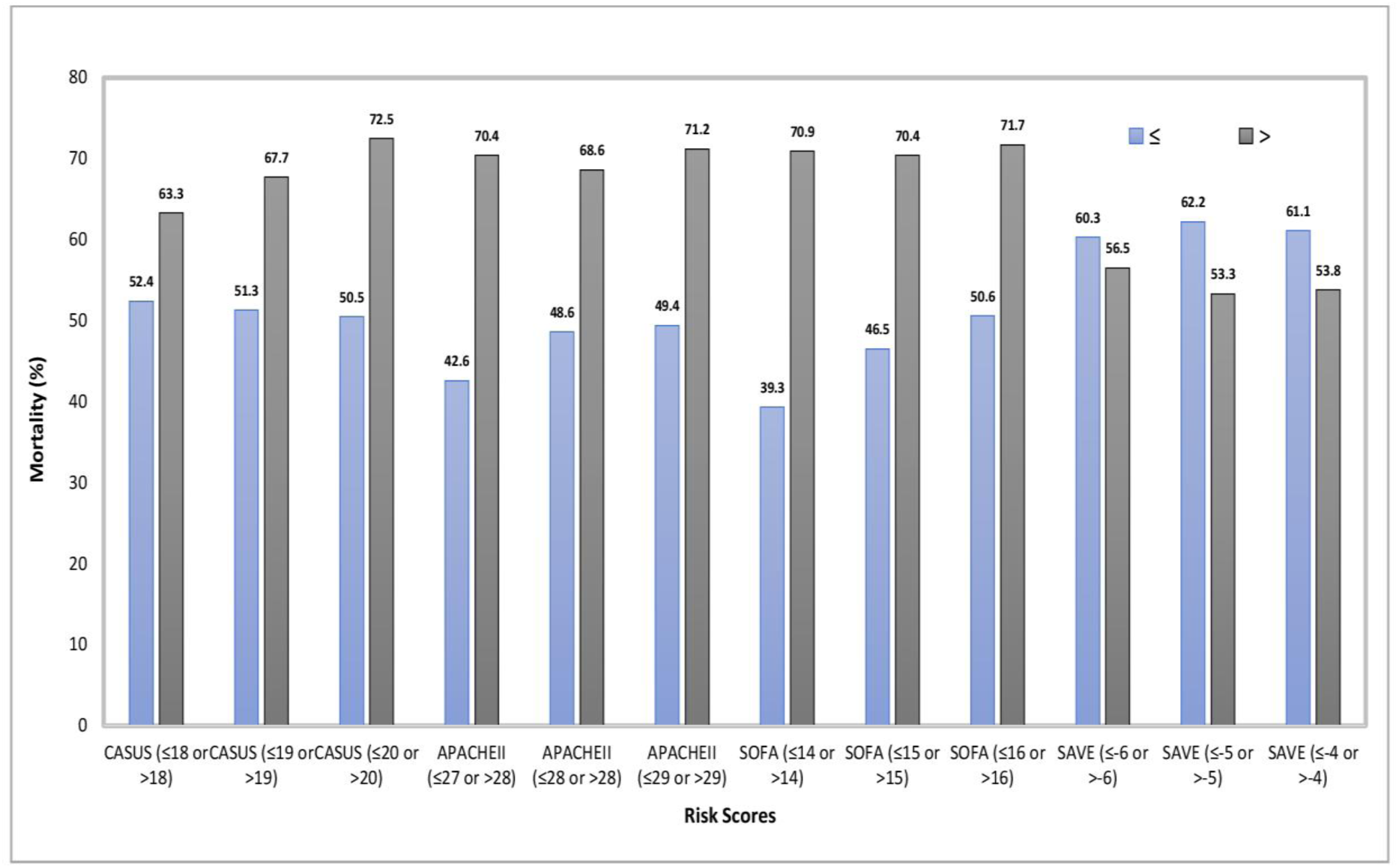
Moratality across different cut-offs for each risk score.

## Discussion

Most of the studies analyzing risk scores have relied on calculation of the score at the initiation of ECMO. We chose not to do so, as these values, if recorded at the initiation of ECMO, would be grossly deranged due to the preceding cardiac arrest or low cardiac output state and may not truly reflect the hemodynamic and organ protection support afforded by MCS. We felt that calculation of risk score at 24 hours of ICU admission after VA-ECMO would track the trend of organ system recovery or the lack of it, thereby providing early insights into the effectiveness of MCS. Predicting a cut-off score at 24 hours of ICU admission for mortality would help clinicians in tailoring therapy appropriately (escalation or de-escalation), re-evaluate management strategies, explore options for definitive treatment of underlying pathology (Percutaneous or surgical revascularization, escalation to durable ventricular assist devices / transplant etc)

The study population was unique compared to those of similar previous studies; the patients were comparatively younger (mean age of non-survivors and survivors was around 50), had a mixed racial profile and had an overwhelming male predominance. (11,16). The survival to discharge in our study was 41.5 %, which parallels the latest published international standards for adult patients undergoing VA-ECMO insertion, derived from Extra Corporeal Life Support Organization (ELSO) registry (46 % for cardiogenic shock and 31 % for ECPR). 84.5% of the patients in our study survived ECLS as compared to 61% in cardiogenic shock and 43 % in ECPR from ELSO registry (17). This is probably because our study group comprised of relatively younger patients and had very few out of hospital cardiac arrests.

Previous studies have identified age, cardiac arrest before ECMO, duration of CPR, serum lactate levels before ECMO institution and duration of ECMO as predictors for survival after VA-ECMO (12–15, 18,19). Our study found that amongst pre-ECMO variables, only serum lactate and serum creatine was significantly associated with mortality. It is interesting to note that, age was not significantly associated with mortality in our study, this could be due to relatively younger population studied, with the mean age amongst survivors and non-survivors being around 50 years.

The APACHE-II and SOFA scores have been used extensively to predict mortality after ICU admissions and have been externally validated (20–23) CASUS was introduced as a specialized cardiac surgery scoring system used for prognostication and mortality prediction after cardiac surgery and has been validated by various studies. (24–26). Several specific predictive scores in VA-ECMO have been proposed, including Survival After Veno-arterial ECMO (SAVE), prEdictioN of Cardiogenic shock OUtcome foR AMI patients salvaGed by VA-ECMO (ENCOURAGE), pRedicting mortality in patients under-going veno-arterial Extracorporeal MEMBrane oxygenation after coronary artEry bypass gRafting (REMEMBER), and modified SAVE scores (12–15). All these scores focused on pre-cannulation variables in predicting mortality after VA-ECMO. All these scores have been derived from selective clinical sub-sets of cardiogenic shock, hence applicability across various clinical scenarios causing cardiogenic shock may be sub-optimal.

SOFA and APACHE-II have been used in predicting outcome in patients on VA-ECMO, despite not being derived or intended to be used for the same. These scores have been externally validated and found to be comparable to specific scores developed for ECMO patients, in terms of discriminatory ability. (27). There have been various studies which found modest discriminatory ability for SOFA and APACHE-II in mortality prediction in patients on VA-ECMO (9,13,28,29), which was similar to the results of our study. All the above-mentioned studies calculated the risk scores at the time of initiation of VA-ECMO, in contrast, our study recorded values, clinical parameters and calculated scores at 24 hours of admission to ICU after initiation of VA-ECMO.

CASUS, developed as a specialized post-cardiac surgery scoring system, has not been extensively used to assess outcome in VA-ECMO, nor has it been externally validated. The thought process behind using CASUS in our study was that since it was developed as a specialized cardiac surgery score, its discriminative ability would be similar in ECLS population. Likewise, Hoffman et al, retrospectively analyzed 90 adults who underwent VA-ECMO insertion and found that CASUS as compared to SOFA, calculated 12 hours after ECMO initiation had better discriminatory ability for mortality prediction (30). Their finding contrasted with that of our study, in which SOFA had better predictability for mortality than CASUS. Larger proportion of post cardiotomy ECMO in the Hoffman study (75%) as compared to ours (29%) could have had a bearing in the contrasting results.

Schmidt et al. created the SAVE scoring system utilizing the ELSO registry to predict survival to discharge in VA-ECMO patients (12). The main drawback of SAVE score was that it did not include patients who underwent ECPR. There have been many studies in which the SAVE score had demonstrated modest discriminatory power in predicting mortality (17,27,30–32). Our study found very poor discriminatory ability for SAVE score (AUC 0.44) and it fared the worst amongst the 4 scores studied. This is probably due to a large proportion of ECPR cases in our study population (50%) and the SAVE score was developed without ECPR patients.

The findings of our study parallel that of various studies which compared different risk scores in patients on VA-ECMO. B. Worku et al retrospectively analyzed fifty-one patients undergoing institution of VA-ECMO support, APACHE II, SOFA, SAPS II (Simplified Acute Physiology II), Encourage, SAVE, and ACEF (age, creatine, ejection fraction) scores were calculated and their ability to predict outcomes were assessed. The results showed modest discriminatory ability of the scores in the cohort of patients and ACEF score performed best (17). Compared to our study, this study had, slightly older patients and had higher proportion of females. Compared to our study, the performance of APACHE-II was similar, SOFA fared worse, and SAVE better. The reason for SAVE score to fare better could be linked to a smaller number of ECPR patients as compared to our study group (41% vs 50%).

Wengenmayer et al. developed PREDICT VA-ECMO based on point-of-care test (POCT) measurement lactate, pH and standard bicarbonate concentration and compared with SAVE, APACHE-II, SOFA and SAPS scores in predicting hospital survival after VA ECMO.PREDICT VA-ECMO score was able to predict hospital survival reliably in the derivation as well as in the validation cohorts and outperformed the comparator scores. The 12-hour PREDICT VA-ECMO integrated lactate, pH and standard bicarbonate concentration at 1 hour, 6 hours and 12 hours after ECMO insertion allowed even better prognostication (33). Unlike most ECMO risk scores which heavily rely on pre-ECMO variables and are calculated at the initiation of ECMO, PREDICT VA-ECMO relied on parameters after initiation of ECMO at regular intervals. This parallels our study methodology where we calculated the risk scores 24 hours after initiation of ECMO. The performance of APACHE-II and SOFA in the study was similar to our findings, but that of SAVE score was higher than that in our study, albeit with similar proportion of ECPR (51% vs 50%). The PREDICT VA-ECMO, relies only on 3 POCT parameters of tissue perfusion to predict mortality. The score fails to address the recovery or lack of it, of major organ systems (CNS, respiratory, renal etc.) which are important aspects in risk prediction and prognostication.

Muller et al formulated the ENCOURGE score in patients undergoing VA-ECMO after acute myocardial infarction (AMI) to predict mortality. Based on multivariable logistic regression analyses, the ENCOURAGE score was constructed with seven pre-ECMO parameters. The ENCOURAGE score performed better than SAVE, SAPS II, and SOFA scores. (14). As compared to ours, in this study SAVE score fared better, with SOFA showing similar predictability Wang et al. proposed REMEMBER score for predicting mortality after VA-ECMO in patients who underwent coronary artery bypass grafting (CABG). The REMEMBER score performed better than the SOFA, SAVE, EUROscore, and ENCOURAGE scores in this population (15). Compared to our study, this study was exclusively on post-cardiotomy ECMO, comprised of older patients, and had lesser proportion of ECPRs. The mortality in this study was comparable to ours (55% vs 58.5%), but successful weaning from ECMO was much reduced (64% vs 84.5%). SOFA and SAVE scores, which were calculated at the time of, initiation of ECMO, fared better than our study based on AUC.

Pladet LCA et al conducted a systematic review and metanalysis of studies focusing on mortality prediction in the setting of ECMO (27). The study found that the discriminatory ability of frequently validated ECMO scores (SAVE, RESP score) were moderate and were comparable to general ICU risk scores (APACHE-II, SOFA, SAPS-II). They found that majority of models had a high risk of bias and were conditional on the fact that ECMO support had already been initiated The findings of our study have been in line with previous studies comparing specific ECMO scores and general ICU scores, where only moderate predictive ability was associated with both general ICU and ECMO specific scores. The reason for the average performance of ECMO specific scores historically, could be due to the fact that majority are calculated at the time of ECMO insertion and hence cannot track the clinical course of the patient on ECMO. Our study found that in combination, the four risk scores studied had a better predictive ability than each one done in isolation. The predictive ability was further enhanced by addition of other specific potentially clinically relevant predictors with non-survivors (duration of ECMO, low flow minutes, Indication for ECMO, location of cannulation, Type of MI associated with cardiac arrest). Another important finding in our study is the association of ECMO flow at 4 hours and 12 hours after ECMO initiation with survival, lower flow rates being significantly associated with survival. Attainment of lower flow rates over a period of time on ECMO signifies recovery of left ventricular function; and as such whenever a patient is placed on ECMO efforts should be made to reduce the flow as early as possible by looking for signs of LV recovery (improvement of pulse pressure, echocardiographic parameters etc.). The poor correlation of mortality prediction in our study between score based on pre-ECMO variables (SAVE) and other scores calculated 24 hours after ICU admission after initiation of ECMO points to importance of tracking organ system recovery on extra corporeal circulation. Serial assessment of parameters pertaining to organ sytem recovery, and its interpretation is paramount in predicting outcome in patients put on VA-ECMO.

### Limitations of the study

This was a single-center retrospective study. The most important limitation of this study was the inability to point out the pre-insertion variables responsible for a favorable outcome which could potentially avoid futile insertions. However, this study points to many immediate post ECMO insertion parameters (some of them modifiable) which may be able to have a positive impact on the overall results. Our study had very few post cardiotomy patients and patients with out of hospital cardiac arrest. Another potential limitation is the moderate AUC of the predictors which mandates future validation studies.

### Conclusions

Our study comparing predictive performance of general ICU, cardiac surgery and ECMO risk scores showed that the general ICU and cardiac surgery score fared better than the ECMO specific score. The APACHE-II, SOFA and CASUS, calculated at 24 hours of ICU admission were significantly higher amongst non-survivors as compared to survivors. APACHE-II, developed for general ICU patients, demonstrated the best mortality predictive ability, although with moderate discriminatory power, in our cohort of patients. APACHE-II scores of 27 or above and SOFA of 14 or above at 24 hours of ICU admission after ECMO cannulation can predict mortality and will aid physicians in decision making. SAVE score is unsuitable in predicting mortality in ECPR. ECMO flow at 4 hours and 12 hours after ECMO initiation can predict mortality on ECMO. Further studies, to develop dynamic scoring systems, focusing on patients’ clinical course on ECMO, and tracking the trend of mortality variables are required to enhance discriminatory power of scores for predicting mortality of VA ECMO accurately.

## Declarations Institutional approval

The study was approved by the ethics committee of the institute (reference number MRC-01-22-701),). The need for informed consent was waived by the institute as no specific intervention was being done, and sampling blood for lab investigations was part of routine post-operative care.

## Consent for publication

The ethical committee of the institute (reference number – MRC −01-22-701), approved to publish this manuscript

## Availability of data and material

The generated datasets from the study or subsequent analysis are not available in the public domain. This can be requested from the corresponding author.

## Authors’ contributions

Suraj Sudarsanan: conceptualization, data curation, investigation, funding acquisition, writing original draft, visualization.

Praveen Sivadasan: data curation, software, review and editing. Prem Chandra: formal analysis, methodology, visualization

Kathy Atuel, Hafeez Lone, Hany Osman and Irshad Ehsan: data curation

Amr Omar, Cornelia Carr, Abdul Rasheed Pattath and Abdulaziz Alkhulaifi: review and editing Yasser Shouman, Abdulwahid Almulla: supervision.

## Data Availability

The generated datasets from the current study are not publicly available but are available from the corresponding author on reasonable request through the regulations of the medical research center of the Institute.

## Acknowledgments

This project would not have been possible without the unlimited help and support of many individuals and our organization. The authors thank all members of the Cardiothoracic surgery department, of the Institute, for providing all required data related to this work. The authors also extend their thankfulness to the medical research department of the Institute for continuous support throughout this project.

## Availability of data and material

All methods used in this study were carried out in accordance with relevant guidelines and regulations (Declaration of Helsinki).

## List of abbreviations

ACEF: Age, Creatine, Ejection Fraction
AKI: Acute kidney injury
AKIN: Acute Kidney Injury Network
AMI: acute myocardial infarction
APACHE II: Acute Physiology and Chronic Health Evaluation II
aPTT: Activated partial thromboplastin time
AUC: Area under the curve
CABG: coronary artery bypass grafting
CASUS: Cardiac Surgery Score
CI: Confidence interval
CPB: Cardio-pulmonary bypass
CRRT: Continuous renal replacement therapy
CS: Cardiogenic shock
CTICU: Cardiothoracic Intensive Care Unit
ECLS: Extracorporeal life support
ECMO: Extra-corporeal Membrane oxygenation
ECPR: Extracorporeal cardio-pulmonary resuscitation
ELSO: Extra Corporeal Life Support Organization
ENCOURAGE: prEdictioN of Cardiogenic shock OUtcome foR AMI patients salvaGed by VA-ECMO
IABP: Intra-aortic balloon pump
ICU: Intensive Care Unit
IQR: inter-quartile range
LR+: likelihood ratio of a positive test
LR-: likelihood ratio of a negative test
LOS ICU: Duration of ICU stay
LOS HOSP: Duration of Hospital stay
LOV: Length of Ventilation
MCS: Mechanical Circulatory support
NPV: Negative predictive value
POD: Post-operative day
POCT: point-of-care test
PPV: positive predictive value
REMEMBER: pRedicting mortality in patients under-going veno-arterial Extracorporeal MEMBrane oxygenation after coronary artEry bypass gRafting
ROC: Receiver operating curve
SAPS II: Simplified Acute Physiology Score II
SAVE: Survival after VA-ECMO
SD: standard deviation
SOFA: Sequential Organ Failure Assessment
VA-ECMO: Veno-arterial extracorporeal membrane oxygenation
VAV-ECMO: Veno-arterio-venous extracorporeal membrane oxygenation

